# Perinatal Risk and Protective Factors in the Development of Diffuse White Matter Abnormality on Term-Equivalent Age MRI in Infants Born Very Preterm

**DOI:** 10.1101/2020.08.01.20166488

**Authors:** Nehal A. Parikh, Puneet Sharma, Lili He, Hailong Li, Mekibib Altaye, Venkata Sita Priyanka Illapani, for the Cincinnati Imaging & Neurodevelopment Early Prediction Study (CINEPS)

**Author notes:** Correspondence: Nehal A. Parikh, DO, MS, Professor of Pediatrics, Cincinnati Children’s Hospital Medical Center, 3333 Burnet Avenue, MLC 4009, Cincinnati, OH, 45229, United States.

## Abstract

**Importance:** Diffuse white matter abnormality (DWMA) is the most common brain MRI finding in very preterm infants and is predictive of neurodevelopmental impairments. However, its etiology remains elusive and identifying perinatal risk and protective factors may allow clinicians to reduce the burden of DWMA.

**Objective:** To identify perinatal clinical factors that are associated with the development of objectively diagnosed DWMA in very preterm infants.

**Design:** A prospective cohort was enrolled between September 2016 and November 2019. Brain MRIs were collected at 39 to 45 weeks postmenstrual age (PMA) to evaluate DWMA volume. A pre-defined list of pertinent maternal characteristics, pregnancy/delivery data, and neonatal ICU data was collected for enrolled patients to identify antecedents of objectively diagnosed DWMA.

**Setting:** Five level III/IV NICUs in the greater Cincinnati, Ohio area.

**Participants:** A population-based sample of 392 very preterm infants born before 33 weeks gestational age.

**Exposure:** Very preterm birth with associated diseases and treatments.

**Main Outcome and Measure:** Objectively diagnosed DWMA volume on brain MRI at term-equivalent age.

**Results:** 377 of the 392 very preterm infants (96%) had high quality MRI data. Mean (SD) gestational age was 29.3 (2.5) weeks. In multivariable linear regression analyses, pneumothorax (p=.027), severe bronchopulmonary dysplasia (BPD) (p=.009), severe retinopathy of prematurity (ROP) (p<0.001), and male sex (p=.041) were associated with increasing volume of DWMA. The following factors were associated with decreased risk of DWMA – dexamethasone for severe BPD (p=.004), duration of caffeine for severe BPD (p = 0.009), and exclusive maternal milk at NICU discharge (p=.049).

**Conclusions and Relevance:** Severe ROP and BPD exhibited the strongest adverse association with the development of DWMA. Caffeine and dexamethasone treatments for infants with severe BPD exhibited a protective effect against development of DWMA. The beneficial association with maternal milk is also a modifiable factor that has clinical implications.

**Key Points:** *Question:* What perinatal clinical factors are independently associated with the development of diffuse white matter abnormality (DWMA) in very preterm infants?

*Findings:* In this prospective cohort study, pneumothorax, severe bronchopulmonary dysplasia (BPD), severe retinopathy of prematurity, and male sex were significantly associated with an increased risk of DWMA. Significant protective factors included dexamethasone for severe BPD, duration of caffeine for severe BPD, and exclusive maternal milk diet at NICU discharge.

*Meaning:* Knowledge of these common and modifiable neonatal diseases and treatments may allow clinicians to reduce the burden of DWMA development through targeted prevention or treatments approaches.

## Introduction

Improvements in the care of preterm infants have resulted in fewer instances of severe injury such as periventricular leukomalacia (PVL) and intraventricular hemorrhage (IVH)^1^]. Even so, the prevalence of neurodevelopmental impairment (NDI) remains unacceptably high^2,3^. Brain MRI studies in preterm infants have identified diffuse, subtle abnormalities in brain maturation and signal abnormality that may be the modern antecedents of NDI^4-8^]. Diffuse excessive high signal intensity (DEHSI) is defined as the presence of higher than normal signal intensity in the developing white matter, as seen on T2-weighted MRI^1^. It is detected in 50-80% of MRI scans of very preterm infants at approximately term-equivalent age^9^. Detailed diffusion MRI studies have identified a complex set of white matter microstructural abnormalities in areas of the brain affected by DEHSI^10,11^. The only post-mortem analysis of infants with DEHSI identified pathology consistent with PVL, such as microgliosis and astrocytosis, in addition to distinct findings such as diffuse vacuolations^12^. It remains unclear if DEHSI represents a milder form of PVL, because of evidence of diffuse injury without necrosis on post-mortem histology, or a distinct abnormality^13,14^.

Despite these pathological radiographic and histologic findings, only a few studies have explored which perinatal clinical factors contribute to the development of DEHSI, in order to further our understanding of this prevalent signal abnormality^1,9,10,15,16^. In one study, surgical ligation of patent ductus arteriosus (PDA) was identified as a potential risk factor for DEHSI^10^. Several other studies could not identify any antecedents of DEHSI; however, these studies were likely underpowered^1,9,15^. Additionally, these negative results may have stemmed from qualitative diagnosis of DEHSI, which is subjective and cannot be made with high reliability^17-19^. This motivated us to develop automated algorithms to objectively quantify DEHSI^20,21^. We refer to DEHSI, when objectively diagnosed, as diffuse white matter abnormality (DWMA). It is defined as the total volume of white matter exhibiting elevated signal intensity (details below) as compared to the surrounding white matter on term T2-weighted MRI^20,21^. In two prior studies we correlated DEHSI and objectively diagnosed DWMA at term with NDI at age 2 and found that DWMA was significantly correlated with cognitive and language development, whereas DEHSI was not^6,8^.

Previously, we identified severe retinopathy of prematurity (ROP) and severe bronchopulmonary dysplasia (BPD) as antecedent risk factors of objectively diagnosed DWMA^16,22^. However, these prior studies lacked sufficient power to examine less common antecedent factors and common pathophysiologic pathways (e.g. inflammation). Our objective was to identify antenatal and neonatal clinical factors that are independently associated with the development of objectively diagnosed DWMA at term in a large geographically-defined cohort of very preterm infants. We hypothesized that several modifiable clinical antecedent factors would be associated with the development of DWMA, as diagnosed on term MRI.

## Designs & Methods

### Subjects

We enrolled a multicenter prospective cohort of 392 very preterm infants from five neonatal intensive care units (NICU) in the greater Cincinnati area. This included all four academic and community level III/IV NICUs in Cincinnati: 1) Cincinnati Children’s Hospital Medical Center (CCHMC), the primary academic referral center for high-risk neonates; 2) University of Cincinnati Medical Center, the primary academic referral center for high-risk pregnancies; 3) Good Samaritan Hospital; 4) St. Elizabeth’s Healthcare; and one community level III NICU in Dayton, Ohio, Kettering Medical Center. All very preterm infants – born at or before 32 weeks gestational age (GA) – who were cared for in one of these NICUs between September 2016 and November 2019 were eligible for inclusion. Infants were excluded if they met any of the following criteria: 1) known chromosomal or congenital anomalies affecting the central nervous system; 2) cyanotic heart disease; or 3) hospitalization and mechanical ventilation with greater than 50% supplemental oxygen at 45 weeks postmenstrual age (PMA). The Cincinnati Children’s Hospital Institutional Review Board approved the study, and the review boards of the other hospitals approved the study based on an established reliance agreement. Written informed consent was given by a parent or guardian of each study infant, after they were given at least 24 hours in which to review the consent and ask questions of the investigators.

Information about MRI data acquisition^23^, objective DWMA quantification^21,24^, and structural MRI scoring^23,25^ are previously described and summarized in the Supplementary Appendix.

### Clinical antecedents

Trained research staff collected a pre-defined list of maternal characteristics, pregnancy/delivery data, and infant data beginning at birth and ending at NICU discharge or study MRI examination, whichever occurred first. All variables were as previously defined (Supplementary Appendix)^16^. We could not examine the effect of surgical ligation for PDA, as this was performed in only three subjects. All brain MRI scans were read by a pediatric neuroradiologist as previously described^23^, using an established neonatal MRI quantification system to derive a global abnormality score^25^.

### Statistical analysis

DWMA volume data was skewed and was thus transformed by taking its cubic root. As described previously^16^, we examined the association between approximately 50 antenatal, intrapartum, and postnatal clinical factors with normalized DWMA volume in bivariate linear regression analyses. Variables that were correlated with DWMA volume (p<0.10) in bivariate analyses were entered into a multivariable linear regression model in a manual backward stepwise fashion, to evaluate their independent association with DWMA. In addition, we used knowledge of prior literature and biological plausibility to guide variable selection. Because postnatal covariates can overshadow antepartum or intrapartum variables that may be causative, we created multivariable regression models in which we ordered clinical factors temporally, so that the earliest occurring factors were entered first and could not be displaced by later occurring covariates^16,26^.

To control for variation in clinical care practices between the five NICUs, we included NICU/Center as a covariate in the final model. Main effects and interactions were evaluated. All analyses were adjusted for postmenstrual age at MRI scan. Two-sided p values <0.05 were considered to indicate statistical significance. We performed all analyses using STATA 16.0 (Stata Corp., College Station, TX).

## Results

Of the original cohort of 392 infants, 15 could not be accurately segmented, eight because of excessive motion artifacts and seven due to moderate-severe ventriculomegaly/brain injury. Therefore, accurate DWMA data was available for 377 infants (96%). The mean (SD) gestational age was 29.31 (2.50) weeks. Table 1 summarizes key baseline antepartum, intrapartum, and postnatal maternal and infant clinical characteristics for our final cohort. In bivariate analyses (as a first step for selecting candidate variables for the multivariate model), adjusting for PMA at MRI scan, several antecedents were associated with normalized DWMA volume (p<.10), including maternal progesterone therapy (p=.053), male sex (p=.069), pneumothorax (p=.030), prophylactic indomethacin (p<.001), cyclooxygenase inhibitor therapy (ibuprofen/indomethacin) for PDA (p=.004), any BPD (p=.086), severe BPD (p=.041), postnatal dexamethasone for BPD (p<.001), duration of caffeine therapy (p=.085), severe ROP (p=.042), surgery requiring general anesthesia (p=.073), white matter abnormality score (p=.002), and NICU/Center (p=.014). Additionally, we identified a significant interaction between postnatal dexamethasone therapy and severe BPD (p<.001) and between duration of caffeine therapy and severe BPD (p=.006).

**Table 1.** **Distributions of important antenatal, intrapartum, and postnatal clinical factors prior to MRI at term-equivalent age in a multicenter cohort of very preterm infants**.

| **Perinatal Clinical Factors\*** | **All Infants (N=377)** |
| --- | --- |
| Maternal diabetes | 44 (11.7%) |
| Histologic chorioamnionitis† | 104 (30.6%) |
| Maternal progesterone therapy | 73 (19.4%) |
| Antenatal steroids (any) | 348 (92.3%) |
| Antenatal magnesium therapy | 316 (83.8%) |
| Gestational age (weeks), median (range) | 29.7 (23.0–32.9) |
| Birth weight (grams), mean (SD) | 1302 (452) |
| Male sex, n(%) | 198 (52.5%) |
| Apgar score <5 at 5 minutes§ | 48 (12.9%) |
| Pneumothorax | 25 (6.6%) |
| Prophylactic indomethacin therapy | 15 (4.0%) |
| Cyclooxygenase inhibitor therapy | 36 (9.6%) |
| Caffeine therapy | 266 (70.6%) |
| Duration of caffeine therapy in infants with severe BPD, median (range) | 62 (0–89) |
| Lowest mean blood pressure in first 24 hours (mm Hg), mean (SD)** | 31.4 (7.4) |
| Transitional hypotension requiring therapy | 15 (4.0%) |
| Surgery for NEC or SIP | 9 (2.4%) |
| Culture positive late onset sepsis | 41 (10.9%) |
| Patent ductus arteriosus | 94 (24.9%) |
| Severe ROP | 18 (4.8%) |
| BPD (any severity) | 154 (40.9%) |
| Severe BPD | 65 (17.2%) |
| Postnatal dexamethasone for severe BPD | 31 (8.2%) |
| Surgery requiring general anesthesia | 42 (11.1%) |
| White matter abnormality score on term MRI, median (range) | 1 (0-15) |
| PMA at MRI scan (weeks), mean (SD) | 42.7 (1.4) |

\*All values are N (%), unless otherwise noted. Abbreviations: BPD – bronchopulmonary dysplasia, NEC – necrotizing enterocolitis, PDA – patent ductus arteriosus, PMA – postmenstrual age, ROP – retinopathy of prematurity, SIP – spontaneous intestinal perforation
†Pathology not performed in 27 infants; §Data missing for 4 infants; \*\*Data missing for 7 infants

In multivariable linear regression analyses, controlling for PMA at MRI scan and NICU/Center, several postnatal variables remained significant in the final model, including infant sex, pneumothorax, severe ROP, severe BPD, postnatal dexamethasone for severe BPD (interaction term), duration of caffeine therapy for severe BPD (interaction term), exclusive maternal breast milk nutrition at NICU discharge, and white matter abnormality score (Table 2). Both severe ROP and severe BPD increased the risk of DWMA. Conversely, postnatal steroid and caffeine therapy for severe BPD reduced the risk of DWMA (Table 2). The addition of gestational age (p=.854) did not change the significance of the other variables in the final model. Replacing our primary outcome variable – DWMA volume normalized by white matter volume – with either DWMA volume normalized by combined white and gray matter volume or uncorrected DWMA volume did not change the significance level or regression parameters of the covariates by more than 5%.

**Table 2.** **Final multivariable linear regression model displaying the coefficients of several clinical antecedents that were associated with the development of diffuse white matter abnormality (DWMA), which was objectively-defined on structural brain MRI at term-equivalent age**.

| **Clinical Antecedent** | **Coefficient (95% CI)\*** | **P Value** |
| --- | --- | --- |
| Pneumothorax | 0.0328 (0.0038, 0.0618) | 0.027 |
| Severe bronchopulmonary dysplasia (BPD) | 0.0570 (0.0141, 0.0999) | 0.009 |
| Dexamethasone for severe BPD | -0.0551 (-0.0928, -0.0174) | 0.004 |
| Duration of caffeine for severe BPD | -0.0008 (-0.0016, -0.0001) | 0.029 |
| Severe retinopathy of prematurity | 0.0659 (0.0305, 0.1014) | <0.001 |
| Exclusive maternal milk at NICU discharge | -0.0237 (-0.0472, -0.0002) | 0.049 |
| White matter abnormality score on term MRI | -0.0047 (-0.0081, -0.0014) | 0.006 |
| Male sex | 0.0151 (0.0006, 0.0296) | 0.041 |
| Postmenstrual age at MRI scan | -0.0266 (-0.0322, -0.0211) | <0.001 |
| Center | -0.0098 (-0.0173, -0.0023) | 0.011 |

\*Normalized volume of DWMA was transformed by taking its cubic root. Therefore, the coefficients represent approximately the cubic root change in DWMA volume for one unit change in the clinical antecedent, while holding the other independent variables constant.

Of the 65 infants with severe BPD, 39 were treated with postnatal dexamethasone, all using the published Dexamethasone A Randomized Trial (DART) protocol, which results in a cumulative dosing of 0.89 mg/kg of dexamethasone administered over 10 days^27^. Nine of these infants received a second course of DART, and one infant received three total courses. The median (range) duration of caffeine therapy was 62 (0-89) days for infants with severe BPD. At NICU discharge or study MRI, whichever came first, 42 (11.1%) were receiving their mother’s breast milk exclusively, 165 (43.8%) were receiving a combination of mother’s milk and preterm infant formula, and the remaining 170 (45.1%) were exclusively receiving infant formula. Replacing exclusive mother’s milk in the model with exclusive formula at discharge resulted in a beta coefficient of .01431 (95% CI: -.00036, .02898; p=.056), which represents a significant trend towards an opposite effect on DWMA volume.

We did not find a significant relationship in bivariate or multivariable analyses between DWMA and any of the following key clinical variables: antenatal corticosteroid therapy, antenatal magnesium therapy, clinical or histologic chorioamnionitis, delayed cord clamping, low 5-minute Apgar score, transitional hypotension, PDA, sepsis, antibiotics, necrotizing enterocolitis, or duration of total parenteral nutrition.

## Discussion

Studying a well-characterized and geographically-defined cohort of very preterm infants, we identified several new independent perinatal clinical factors that antecede the development of DWMA at term-equivalent age. We also externally validated the previously-reported association between both severe ROP and severe BPD and increased DWMA volume^16^. These two diseases exhibited the largest effect on DWMA volume. Interestingly, postnatal dexamethasone therapy and caffeine therapy, the two most effective drugs for reducing BPD risk, were associated with a decreased DWMA risk when administered to very preterm infants with severe BPD. These novel associations have not been reported previously. They further strengthen the adverse association between severe BPD and DWMA. Pneumothorax and an exclusive diet of maternal milk represent two additional factors that have not been previously reported. Each of the new antecedents we uncovered has been associated with NDI, but the way in which they are mediated by early brain development/injury has not been fully delineated.

This is the third study to identify severe ROP as a key risk factor for DWMA^16,22^. Development of severe ROP is among the most prominent neonatal risk factors for NDI, independent of structural injury on cranial ultrasound^28,29^. Even qualitative structural MRI studies have not been able to mechanistically explain this association. Recently, we identified a negative association between ROP and automatically-quantified sulcal depth, a measure of brain maturation, on term MRI^30^. The association with DWMA suggests an additional mechanism by which ROP is causally associated with NDI^28,29^.

Similar to severe ROP, we previously reported a robust association between severe BPD and DWMA in a separate independent cohort^16^. Unlike this prior report, which was likely underpowered for less common conditions/treatments, in our current study we identified an interaction between severe BPD and postnatal dexamethasone therapy and an interaction between severe BPD and duration of caffeine therapy, both of which were associated with reduced DWMA volume. These findings are highly relevant for the clinical care of very preterm infants as they contribute to our understanding of how these drugs may improve long-term neurodevelopmental outcomes, particularly when administered to infants at high risk for severe BPD. For example, postnatal dexamethasone use, even at the relatively low doses used in our population (e.g. DART protocol^27^), has remained low since 2003, when the American Academy of Pediatrics recommended a moratorium due to concerns regarding NDI following dexamethasone therapy^31^. However, it soon become clear that NDI occurred primarily in treated infants at low baseline risk of BPD (<50%), typically during the first week of age, when such risk is difficult to estimate^32^. Conversely, treatment of infants at high baseline risk of BPD (>50-67%) results in a lower risk of cerebral palsy or death. Our finding of an association between dexamethasone treatment in infants with severe BPD and DWMA development suggests a beneficial mechanism of this corticosteroid. Dexamethasone may act on the brain either directly via an anti-inflammatory effect (e.g. reduced cerebral edema) or indirectly by reducing BPD severity. Our novel findings supports the use of low dose dexamethasone for the prevention of DWMA and potentially NDI in infants with severe BPD.

The association of DWMA with duration of caffeine therapy in infants with severe BPD suggests that the proven long-term beneficial effects of caffeine on motor development^33^ in very preterm infants may be mediated through an early neuroprotective effect on DWMA development. Mechanistically, this benefit may be directly mediated by caffeine’s weak diuretic effects, via protection against intermittent hypoxemia^34^, and/or through adenosine receptor blockage and secondary anti-inflammatory effects^35,36^. Conversely, it may act indirectly by reducing the risk of BPD^37^. Our novel findings suggest that DWMA volume could potentially be used as a surrogate outcome measure in quality improvement efforts or new randomized trials, in order to assess the expanded use of new therapeutic dosing of caffeine or dexamethasone in infants with severe BPD.

The cognitive benefits of human milk in preterm infants were demonstrated in a meta-analysis of 11 cohort studies with short-term outcomes^38^ and in studies of preterm infants at school age and in adolescence^39-41^. We identified a significant protective effect of an exclusive maternal milk diet during the NICU period on DWMA development at term. Conversely, an exclusive formula diet at NICU discharge exhibited a trend towards higher DWMA volume, and a mixed human milk and formula diet exhibited an intermediate effect (p=0.49). These findings have not been reported previously and add to our growing understanding of the neuronal mechanisms by which maternal milk confers neurocognitive benefits. In quantitative MRI studies, Pogribna et al.^42^ identified dose-dependent improved microstructural maturation of the corpus callosum at term in breast milk-fed infants, while Belsa et al.^43^ reported significant benefits on the corpus callosum and several other white matter regions, including the periventricular white matter, in a smaller cohort of very preterm infants. Other studies have noted increased regional brain volumes, such as deep nuclear gray matter at term^40^ and total white matter in adolescence, in subjects who were predominantly fed maternal milk during the NICU period^41^.

No prior studies have identified an association between neonatal pneumothorax and the development of DWMA. However, several studies in preterm infants have identified an association between pneumothorax and an increased risk of severe IVH^44-46^. In a cohort of 803 very low birth weight infants, pneumothorax was significantly associated with lower odds of survival without severe IVH and/or PVL^46^. While the link between severe IVH on ultrasound and neurodevelopmental deficits is well-established, one large cohort study of 1749 extremely low birth weight infants with normal head ultrasound also identified an independent relationship between pneumothorax and cerebral palsy, suggesting other mechanisms by which pneumothorax adversely affects neurodevelopment^47^. Acute pneumothorax results in acute hypoxemia and hypercapnia in newborns, and in experimental models it can cause cytotoxic cerebral edema^48^. The presence of a tension pneumothorax additionally compromises cardiac output, which limits the hypoxia/hypercarbia-mediated increase in perfusion to the brain and heart^49^. Less severe cases of pneumothorax that do not result in tension and/or cardiovascular compromise may still result in neuronal injury and secondary edema that manifest as diffuse excessive high signal intensity abnormality on term MRI.

The bright signal abnormalities that are the hallmark of DWMA begin to decrease between 42 and 50 weeks PMA^50^. We found a similar pattern of decrease in DWMA with advancing age at MRI scan. Interestingly, we also observed a negative association between objectively-defined DWMA and a semi-quantitative measure of WM injury/abnormality. This association has not been previously reported and suggests that other white matter abnormalities/injuries may result from factors different that those associated with DWMA. Importantly, as with white matter abnormalities such as PVL and ventriculomegaly, objectively-defined DWMA is predictive of cognitive and language deficits, a finding that has now been reported in two independent cohorts of preterm infants^6,8^.

Both ROP and BPD share systemic inflammation, hypoxia, and hyperoxia as common mechanisms that could directly or indirectly result in brain injury. In addition to the well-known risk factors of hypoxia and hyperoxia, recent evidence from large epidemiologic investigations and experimental models implicate antenatal inflammation and postnatal infection/inflammation as strong risk factors for the development of ROP^51-55^. The large reduction in BPD incidence noted in randomized trials of postnatal corticosteroids and the lack of benefit observed following antioxidant therapies or tighter control of oxygen saturation, suggests that inflammation may be a significant causal factor in the development of BPD^56-60^. Our findings of reduced DWMA following treatment with dexamethasone, a potent anti-inflammatory drug, and caffeine^35,36^, further supports inflammation as a key mechanism in the development of DWMA. Conversely, we did not observe an association between DWMA and other pro-inflammatory conditions such as chorioamnionitis or postnatal sepsis. Human milk also has anti-inflammatory properties, as it contains molecules such as secretory immunoglobulin A, transforming growth factor beta, lactoferrin, and interleukin-10 that can protect the newborn against inflammation^61,62^. We also did not find an association with delayed cord clamping, low Apgar scores, transitional hypotension, or PDA, factors more closely associated with hypoxia-ischemia, suggesting that this may not be the predominant underlying mechanism in the development of DWMA.

The strengths of our study include a robust, geographically-based cohort, the use of an objective, validated algorithm to diagnose DWMA, and comprehensive examination of important perinatal variables known to be associated with NDI. Our study also has several weaknesses. We did not collect serum biomarkers of inflammation, hypoxia, or ischemia to further elucidate the molecular mechanisms underlying DWMA development. The rates of PDA ligation and surgery for NEC or SIP were low, and therefore we may have been underpowered to examine their associations with DWMA. Finally, our findings should only be viewed as associations rather than primary causes. Nevertheless, this study represents the largest and most rigorous examination of the antecedents of DWMA development in very preterm infants. The significant relationship between these novel antecedent factors and DWMA enhances our understanding of the mechanisms of their known adverse/protective effects on NDI. It further suggests that quality improvement and research interventions that target reduced rates of ROP, pneumothorax, and BPD, e.g. with increased targeted use of dexamethasone and/or caffeine, may result in a lower burden of DWMA and subsequently in reduced rates of NDI.

## Data Availability

The datasets generated during and/or analyzed during the current study are available from the corresponding author on reasonable request.

## Acknowledgements

This research was supported by National Institutes of Health grants R01-NS094200-05 and R01-NS096037-03 from the National Institute of Neurological Disorders and Stroke (NINDS). The content is solely the responsibility of the authors and does not necessarily represent the official views of NINDS or the National Institutes of Health. We sincerely thank the Cincinnati Imaging & Neurodevelopment Early Prediction Study (CINEPS) Investigators: Principal Investigator: Nehal A. Parikh, D.O., MS. Collaborators (in alphabetical order): Mekibib Altaye, Ph.D., Anita Arnsperger, RRT, Traci Beiersdorfer, RN BSN, Kaley Bridgewater, RT(M.R.) CNMT, Tanya Cahill, MD, Kim Cecil, Ph.D., Kent Dietrich, RT, Christen Distler, BSN RNC-NIC, Juanita Dudley, RN BSN, Brianne Georg, BS, Cathy Grisby, RN BSN CCRC, Lacey Haas, RT(M.R.) CNMT, Karen Harpster, Ph.D., OT/RL, Lili He, Ph.D., Scott K. Holland, Ph.D., V.S. Priyanka Illapani, MS, Kristin Kirker, CRC, Julia E. Kline, Ph.D., Beth M. Kline-Fath, MD, Hailong Li, Ph.D., Matt Lanier, RT(M.R.) R.T. (R), Stephanie L. Merhar, MD MS, Greg Muthig, BS, Brenda B. Poindexter, MD MS, David Russell, JD, Kari Tepe, BSN RNC-NIC, Leanne Tamm, Ph.D., Julia Thompson, RN BSN, Jean A. Tkach, Ph.D., Sara Stacey, BA, Jinghua Wang, Ph.D., Brynne Williams, RT(M.R.) CNMT, Kelsey Wineland, RT(M.R.) CNMT, Sandra Wuertz, RN BSN CCRP, Donna Wuest, AS, Weihong Yuan, Ph.D. We also greatly appreciate the support of our NICU fellows, nurses, and staff, and, most importantly, all the study families that made this research possible.

---

**Figure 1.**
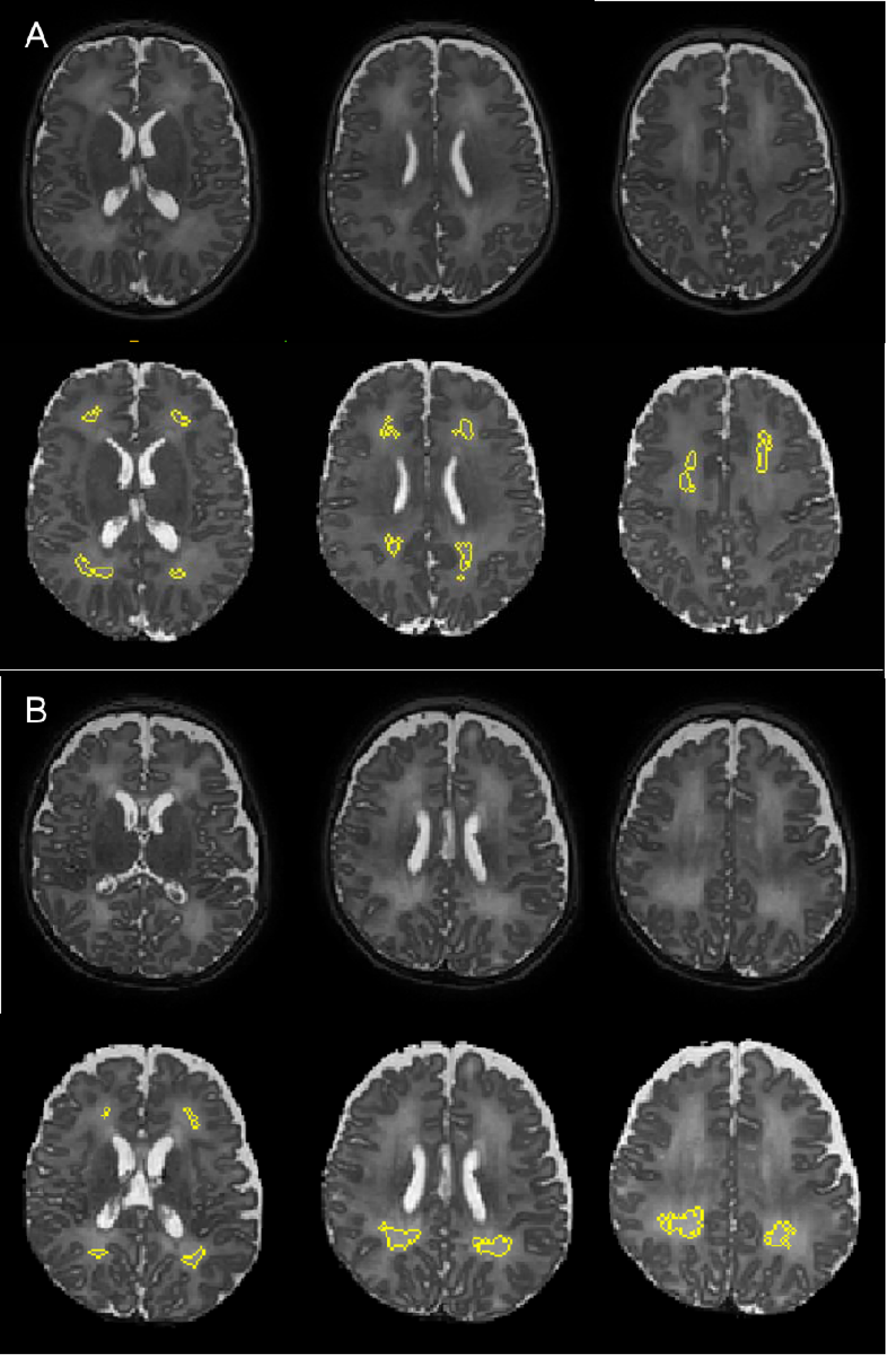
Semiautomated segmentation of diffuse white matter abnormality (DWMA) in the periventricular and central white matter. A. Top three panels display raw axial T2-weighed magnetic resonance images through the central part of the brain from a 28 week very preterm boy. Higher signal intensity can be appreciated in the white matter from the surrounding gray matter. Bottom three panels display the segmented (yellow) moderate degree of DWMA white matter regions. B. Top three panels display raw axial T2-weighed MR images from a 30 week very preterm boy. Higher signal intensity can be appreciated in the periventricular and central white matter from the surrounding gray matter. Bottom three panels display the segmented (yellow) severe DWMA.

